# Retrospective Methodology to Estimate Daily Infections from Deaths (REMEDID) in COVID-19: the Spain case study

**DOI:** 10.1101/2020.06.22.20136960

**Authors:** David García-García, María Isabel Vigo, Eva S. Fonfría, Zaida Herrador, Miriam Navarro, Cesar Bordehore

## Abstract

The number of new daily infections is one of the main parameters to understand the dynamics of an epidemic. During the COVID-19 pandemic in 2020, however, such information has been underestimated. Here, we propose a retrospective methodology to estimate daily infections from daily deaths, because those are usually more accurately documented. This methodology is applied to Spain and its 19 administrative regions. Our results showed that probable infections during the first wave were between 35 and 42 times more than those officially documented on 14 March, when the national government decreed a national lockdown and 9 times more than those documented by the updated version of the official data. The national lockdown had a strong effect on the growth rate of virus transmission, which began to decrease immediately. Finally, the first inferred infection in Spain is about 43 days before it was officially reported during the first wave. The current official data show delays of 15-30 days in the first infection relative to the inferred infections in 63% of the regions. In summary, we propose a methodology that allows reinterpretation of official daily infections, improving data accuracy in infection magnitude and dates because it assimilates valuable information from the National Seroprevalence Studies.

## 1. Introduction

The key parameter to understand and model the evolution of the COVID-19 pandemic is the number of daily infections. Despite its importance, however, it is difficult to estimate reliable data due to the bias in official figures^1^. For symptomatic patients, there are delays of 5.79 days from infection to symptom onset and 5.82 days from symptom onset to diagnosis^2^. In addition, the presence of asymptomatic people and those with mild symptoms hinder their detection. For example, 86% of infections were undetected in Wuhan, China, prior to 23 January 2020, when travel restrictions were imposed^3^. Additionally, not all patients with COVID-19 compatible symptoms are tested, especially at the beginning of the pandemic. So, it is widely assumed that the reported infections are just a fraction of the actual ones. In Spain, a National Seroprevalence Study based on ∼55,000 random participants estimated that only 10.7% (CI 95%: 10.1%-11.3%) of the actual infections had been detected during the first wave (before 22 June 2020) in Spain^4, 5^.

Another problem is the inconsistency of daily infection time series and the variability of the case definitions. For example, on 13 January 2020, China changed the parameters to confirm new cases and ∼15,000 new infections were counted in a single day, while daily infections during the previous week were less than 3,500^6^. In Spain, the method of counting official numbers of infections has been modified several times since the pandemic was decreed. On 22 April 2020, a total of 213,024 cases were reported from positive PCR and antibody tests^7^; however, total cases decreased to 202,990 the next day and thereafter, because only positive PCRs were included in the total cases^8^. Accumulated cases on a given day are obtained by adding the new cases that day to the accumulated cases of the previous day. That has not always been the method, however, because some Spanish regions reviewed and modified the cases in previous days^9, 10^. This inconsistency of time series hampers any kind of accurate analysis. Additionally, data are continuously being updated. In Spain, official daily infection data available during the first wave reported 1,832 new infected on 14 March 2020, but current official data (as downloaded in February 2021) reported 7,478 new cases for the same date.

High fidelity time series of each parameter of an epidemic are crucial to run reliable epidemiological models. The number of new infections per unit time is one of the essential parameters. The official data do not reflect the actual day of infection nor the real number of infections. To overcome this limitation, we propose a retrospective methodology to infer daily infections from daily deaths, believing the number of deaths to be a more reliable parameter than the official number of daily infections^1^. These estimated infections are closer to the date of infection, avoiding delays from the incubation period and symptom onset to diagnosis. Then, the time series of daily infections would help to better understand the pandemic dynamics and to quantify the effectiveness of the different control strategies, avoiding the bias of the official data. In addition, those numbers can be used to feed epidemiological models with more realistic data. Finally, we will apply this methodology in Spain as a whole and each of its 19 administrative regions (17 autonomous communities and 2 autonomous cities).

## 2. Methods

### 2.1 REMEDID

We named this methodology REMEDID, **RE**trospective **M**ethodology to **E**stimate **D**aily **I**nfections from **D**eaths and it is described as follows. Date of infection (*DI*) of an individual may be estimated from the death date (*DD*) by subtracting the incubation period (*IP*) and illness onset to death (*IOD*) periods, hence

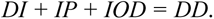

Therefore, *DI* can be estimated from *DD* as far as *IP* and *DD* are known; however, neither *IP* nor *IOD* are fixed values. On the contrary, they are random variables that can be approximated by probability distributions. From dozens of cases in Wuhan, Linton et al.^11^ approximated *IP* for COVID-19 with a lognormal distribution, *X_IP_*, with a mean of 5.6 days and median of 5 days, and *IOD* with a lognormal distribution, *X_IOD_*, with a mean of 14.5 days and median of 13.2 days. Therefore, the infection to death period is a random variable, *X_IP+IOD_*, that follows the distribution *X_IP_* + *X_IOD_*.

Although the addition of two lognormal distributions does not follow any commonly used probability distribution, its probability density function (PDF) can be estimated convolving the PDF of the two variables^12^. If *g(t)* and *h(t)* are the PDF of *IP* and *IOD*, respectively, then their convolution defines the PDF of *X_IP+IOD_*,

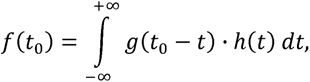

where *t_0_* is a positive real number representing days from infection. Then, *f(t)* is the PDF of the time from infection to death, with a mean of 20.1 days and a median of 18.8 days. Figure 1 shows *g(t)*, *h(t)*, and *f(t)*, as well as a lognormal PDF with the same mean and median as *f(t)* for comparison. Note that the probability that *X_IP+IOD_* is less than or equal to 33 days is 0.95.

**Figure 1.**
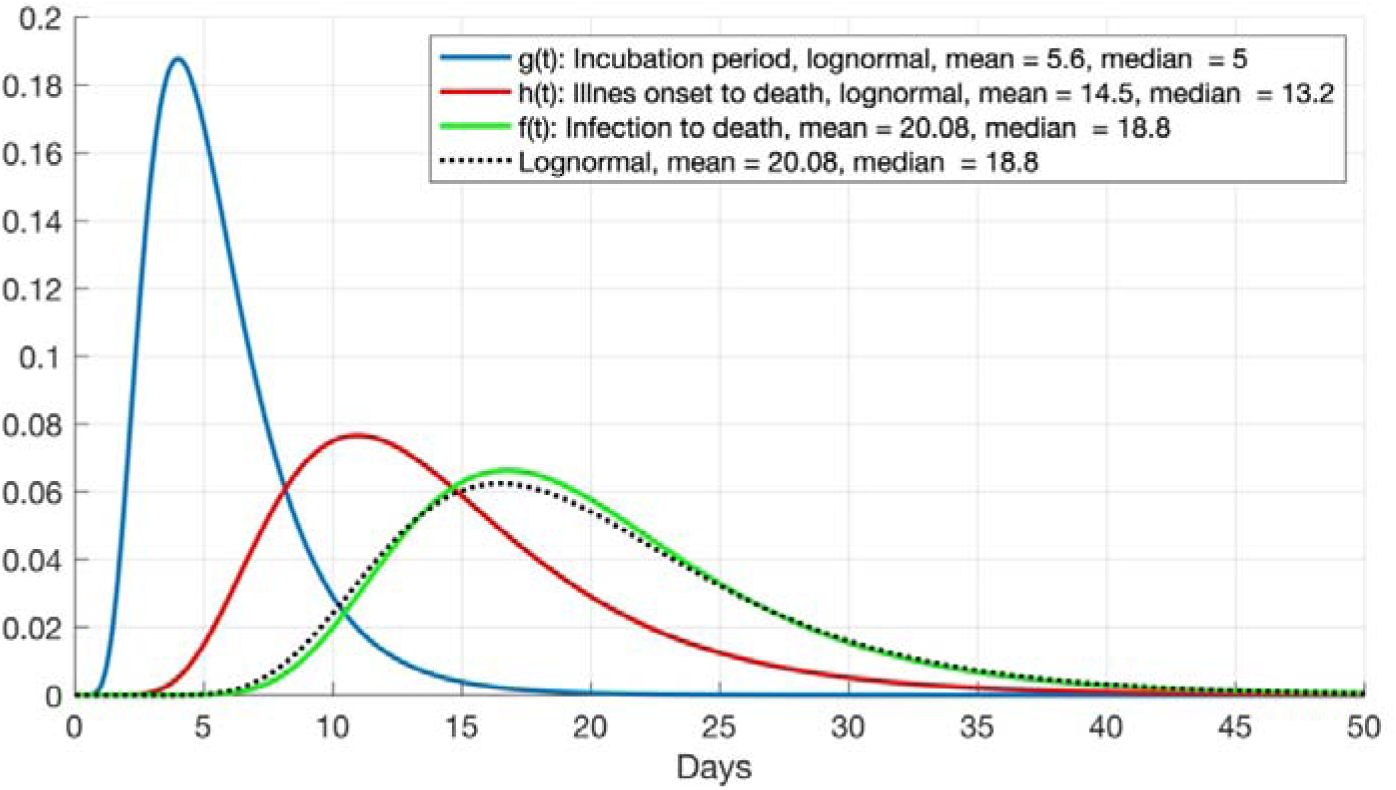
Probability density functions (PDF) of incubation period (*IP*, blue line) and illness onset to death period (*IOD*, red line) from Linton et al. (2020). Green line, *f(t)*, is the convolution of both and represents the PDF of infection to death. Black dotted line is a lognormal PDF with the same mean and median as *f(t)*.

Given a time series of deaths produced by the illness, *x(t)*, we estimate the infection time series that produced such deaths, *y(t)*. If we assume a case fatality ratio (CFR) of 100%, *y(t)* would represent all daily infections. To calculate *y(t)*, we use the following likelihood-based estimation procedure.

Because the relative likelihood that a given infection will produce a death *t* days later is *f(t)*, the infections at a given time *t_0_* can be estimated as

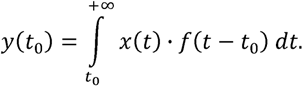

In practice, *x(t)* is a discrete time series and could be written as *x(n)*, where *n* is an integer representing entire days. Let *F(n)* be a discrete approximation to *f(t)* as follows:

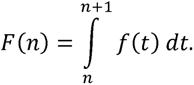

Then,

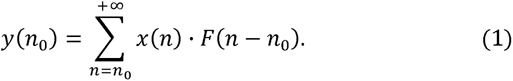

A similar analysis was used previously to estimate a time-variable effective reproduction number for the severe acute respiratory syndrome (SARS) in 2003^13^. For a different CFR, the associated/estimated infections are estimated from *y(n)* in Equation 1 as

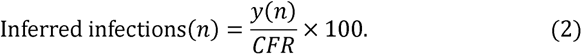

Equations 1 and 2 constitute the kernel of the REMEDID algorithm. Validation of the capacity of REMEDID to reproduce the number of new daily infections is in Appendix A.

### 2.2. Data

#### 2.2.a. Official numbers of COVID-19 infections and deaths

COVID-19 daily infections and deaths for Spain are reported by the *Centro de Coordinación de Alertas y Emergencias Sanitarias* (CCAES), which depends on the Ministry of Health, Social Services and Equality. Time series for the 19 regions of Spain can be downloaded from the national organization that compiles and publishes the data from all regions, the Instituto de Salud Carlos III (ISCIII; https://covid19.isciii.es/). For consistency, time series for Spain were estimated by adding the time series of the 19 regions. The final access to infections from official data was on 16 February 2021 (hereafter referred as *IO_21_*). Those data have been greatly improved with respect to the daily situation reports published by CCAES during the first wave in the first semester of 2020 (hereafter referred as *IO_20_*; https://www.mscbs.gob.es/profesionales/saludPublica/ccayes/alertasActual/nCov-China/situacionActual.htm). For example, *IO_20_* cases were given the date of diagnosis, whereas *IO_21_* cases were given the date at symptoms onset. When the latter was not available, it was estimated as follows: infections prior to 10 May 2020 were given the diagnosis date minus 6 days, whereas since 11 May 2020 only 3 days were subtracted from the date of diagnosis. Asymptomatic cases were given the diagnosis date. These corrections brought reported cases closer to the real infection dates, although they were still delayed by the length of the incubation period. In addition, re-evaluation of the suspicious case of the first infectee on 20 February 2020 in *IO_20_* gave it the date of 1 January 2020 in *IO_21_*. The first official death was reported on 13 February 2020. However, no deaths were reported until 3 March 2020 and afterwards deaths were continuously reported. Then, time spans we considered for *IO_21_* is from 1 January 2020 to 29 November 2020 and for official daily deaths from 3 March 2020 to 1 January 2021, specifically with 33 extra days as required to apply the REMEDID methodology.

When applying REMEDID the use of a reliable CFR is essential. Thus, to determine a realistic CFR in Spain, we used the total infections data from the longitudinal seroprevalence study conducted by ISCIII in 4 phases. The first three phases were completed from 27 April to 22 June and the results were published on 6 July 2020^4, 5^. In this longitudinal cohort study, ∼68,000 people were randomly selected and ∼55,000 of them agreed to be interviewed and tested in the three phases. The study found that 5.2% (CI 95%: 4.9%-5.5%) of the participants presented SARS-Cov2 IgG antibodies.

Extrapolation of this result to the ∼47 million of the population in Spain yielded 2.45 million infections until 22 June, which, with 29,674 accumulated COVID-19 deaths on 22 June, would give a CFR of 1.21% (CI 95%: 1.15%-1.29%). We assumed this value in our analysis for Spain until 22 June. The fourth phase of the study was completed from 16 to 29 November 2020 and the results published on 15 December 2020^14^. In that study, ∼51,000 people agreed to participate. The study found that 9.9% (CI 95%: 9.4%-10.4%) of the participants presented SARS-Cov2 IgG antibodies at any of the four phases. Then, there were 4.75 millions of accumulated infections in Spain, meaning 2.30 million new infections from 23 June to 29 November. Because there were 47,113 accumulated COVID-19 deaths on 29 November (17,439 new deaths from 23 June), it would give a CFR of 0.76% (CI 95%: 0.71%-0.81%). Thus, we assumed different CFRs for the different periods, in order to smooth the discontinuity effects, we propose a CFR time series segmentation as follows: i) a constant value of 1.21% from 1 January to 22 June 2020, ii) a constant value of 0.76% from 30 June to 29 November 2020, and iii) a linear function from 23 June to 28 June that would correspond to the interpolation of the CFR values for 22 June and 29 June. The CFRs of each region were estimated analogously (summarized in Tables 1 and 2). Prior to 22 June, Asturias, Castilla – La Mancha, Castilla y León, Extremadura, País Vasco, and La Rioja had CFRs greater than 1.5%. In contrast, Andalucía, Canarias, Melilla, and Murcia had CFRs less than 0.7%. After 22 June, the CFR for Spain diminished in 0.45%, probably due to improved medical treatments and experience gained during the first wave. In this period, the number of regions with a CFR below 0.7% increased to 10 and those with a CFR greater than 1.5% decreased to 3. Most of the regions revealed certain improvement, particularly remarkable was La Rioja, with a CFR decrease from 3.13% to 1.12%.

#### 2.2.b. Excess of expected deaths from MoMo

Expected deaths from any cause, with an error band representing a 99% confidence interval, are modelled by the Mortality Monitoring (MoMo) surveillance system for 22 countries in Europe (www.euromomo.eu). In Spain, MoMo depends on the ISCIII^15^ and data are available at https://momo.isciii.es/public/momo/dashboard/momo_dashboard.html#nacional. MoMo also collects daily deaths from any cause from the Spanish *Instituto Nacional de Estadística* (INE) and the notaries and civil registries. Daily deaths within the error estimates of the expected deaths are considered usual. Any deviation allows identification of unusual mortality patterns, as for the COVID-19 pandemic. The useful variable here is the excess deaths (*ED*), estimated as the difference between actual and expected deaths. The error band for *ED* can be estimated from the error band of the expected deaths by subtracting the latter from the actual deaths. Note that *ED* can show negative values (Figure 2). By cautiously assuming that all significant *ED* is caused by COVID-19, *ED* can be used to estimate non-reported COVID-19 deaths. In this case, negative values for *ED* make no sense and are set to zero. The MoMo time series was downloaded on 16 February 2021 and we considered the period from 3 March 2020 (first non-isolated official COVID-19 death) to 1 January 2021.

Given that MoMo *ED* in Spain was 45,119 (CI 99%: 37,593-55,098) deaths from 3 March to 22 June, and 23,612 (CI 99%: 11,356-39,690) from 23 June to 29 November, the CFR for the country estimated similarly as in Section 2.1 was 1.85% (CI 95%: 1.74%-1.96%) prior to 22 June, and 1.02% (CI 95%: 0.97%-1.09%) after 23 June. For each region, the CFRs were estimated similarly and are provided in Tables 1 and 2. In all cases, we used a piece wise CFR time series segmented as described in Section 2.2a.

## 3. Results

### 3.1. COVID-19 vs. MoMo

The COVID-19 official deaths and MoMo *ED* time series overlaped for the period from 3 March 2020 to 1 January 2021 for Spain and its 19 regions (Figure 2). In general, there was good agreement between both datasets, meaning that most of MoMo *ED* were related to COVID-19 deaths. During the first wave, the most important differences were observed in Spain, Madrid, Cataluña, Castilla-La Mancha, and Castilla y León. Before 22 June in Spain, MoMo *ED* showed 15,445 accumulated deaths more than the official COVID-19 deaths, which is beyond the error band. That difference comes basically from the four regions with the largest numbers of deaths (Madrid, Cataluña, Castilla-La Mancha, and Castilla y León). Table 1 shows the accumulated values before 22 June, which were used to estimate the CFR for Spain and its 19 regions according to the third phase of the National Seroprevalence Study^4, 5^. For all regions, the CFR estimated from MoMo *ED* was larger than the CFR estimated from COVID-19 deaths. In particular, Asturias, Canarias, and Murcia were twice as large. Ceuta and Melilla dramatically increased their CFR from MoMo *ED*, although that may be biased due to their small populations and numbers of deaths.

Similarly, the same variables for the period from 23 June to 29 November 2020 are reported in Table 2. In Spain, MoMo *ED* showed 6,173 accumulated deaths more than the official COVID-19 deaths. This difference is a third of the difference observed prior to 22 June; because this is within the error band, there was a significant improvement in the detection of COVID-19 deaths in this period. Figure 2 also shows a general agreement between MoMo *ED* and official COVID-19 deaths time series after the first wave, with the exception of late July and early August. These differences were due to two heat waves that were responsible for at least 25% of the MoMo *ED*^16^.

**Table 1.** Accumulated COVID-19 deaths and MoMo Excess Deaths (*ED*) for Spain and its 19 regions. Values from 3 March to 22 June 2020. Population data from INE. *: differences beyond the error estimation.

| Region | 2019<br>Population | Percentage of<br>infections<sup>4,5</sup> | Official data |  | MoMo data |  |  | *ED* minus<br>COVID-19<br>deaths |
| --- | --- | --- | --- | --- | --- | --- | --- | --- |
|  |  | % (CI 95%) | COVID-19<br>deaths | CFR<br>(CI 99%) | *ED* (CI 95%). | Days<br>with<br>lower<br>limit >0 | CFR<br>(CI 95%) |  |
| Spain | 47,026,208 | 5.2 (4.9; 5.5) | 29674 | 1.21 (1.15; 1.29) | 45,119 (37,593;<br>55,098) | 57 | 1.85 (1.74; 1.96) | 15445* |
| Andalucía | 8,414,240 | 3.0 (2.6; 3.6) | 1441 | 0.57 (0.48; 0.66) | 2,123 (651; 5,168) | 23 | 0.84 (0.70; 0.97) | 682 |
| Aragón | 1,319,291 | 4.8 (3.7; 6.2) | 915 | 1.44 (1.12; 1.87) | 1,073 (408; 2,165) | 30 | 1.69 (1.31; 2.20) | 158 |
| Asturias | 1,022,800 | 1.9 (1.3; 2.6) | 336 | 1.73 (1.26; 2.53) | 689 (73; 1,885) | 14 | 3.54 (2.59; 5.18) | 353 |
| Islas<br>Baleares | 1,149,460 | 1.4 (0.8; 2.4) | 226 | 1.4 (0.82; 2.46) | 279 (11; 1,090) | 6 | 1.73 (1.01; 3.03) | 53 |
| Canarias | 2,153,389 | 2.3 (1.5; 3.5) | 166 | 0.33 (0.22; 0.51) | 337 (13; 1,585) | 5 | 0.68 (0.45; 1.04) | 171 |
| Cantabria | 581,078 | 3.7 (2.5; 5.4) | 212 | 0.99 (0.68; 1.46) | 296 (34; 932) | 10 | 1.38 (0.94; 2.04) | 84 |
| Castilla - La<br>Mancha | 2,032,863 | 9.6 (7.8; 11.6) | 2951 | 1.51 (1.25; 1.86) | 5,468 (4,464; 7,003) | 47 | 2.80 (2.32; 3.45) | 2517* |
| Castilla y<br>León | 2,399,548 | 7.8 (6.8; 9.0) | 2829 | 1.51 (1.31; 1.73) | 3,813 (2,763; 5,452) | 45 | 2.04 (1.77; 2.34) | 984 |
| Cataluña | 7,675,217 | 5.9 (5.0; 7.1) | 6507 | 1.44 (1.19; 1.7) | 12,182 (10,034;<br>15,537) | 54 | 2.69 (2.24; 3.17) | 5675* |
| Ceuta | 84,777 | 0.7 (0.3; 1.8) | 5 | 0.84 (0.33; 1.97) | 85 (5; 170) | 3 | 14.24 (5.54;<br>33.22) | 80 |
| C.<br>Valenciana | 5,003,769 | 2.4 (1.9; 3.1) | 1450 | 1.21 (0.94; 1.53) | 2,078 (753; 4,391) | 29 | 1.73 (1.34; 2.19) | 628 |
| Extremadura | 1,067,710 | 3.1 (2.2; 4.3) | 513 | 1.55 (1.12; 2.18) | 919 (298; 2,044) | 25 | 2.78 (2.00; 3.91) | 406 |
| Galicia | 2,699,499 | 1.9 (1.5; 2.4) | 618 | 1.20 (0.95; 1.53) | 755 (122; 2,472) | 13 | 1.47 (1.16; 1.86) | 137 |
| Madrid | 6,663,394 | 11.7 (.3; 13.3) | 8720 | 1.12 (0.98; 1.27) | 14,443 (12,766;<br>16,612) | 62 | 1.85 (1.63; 2.10) | 5723* |
| Melilla | 86,487 | 3.4 (2.3; 5.1) | 2 | 0.07 (0.05; 0.10) | 63 (0; 144) | 0 | 2.14 (1.43; 3.17) | 61 |
| Murcia | 1,493,898 | 1.6 (1.0; 2.7) | 147 | 0.62 (0.36; 0.98) | 387 (37; 1,437) | 9 | 1.62 (0.96; 2.59) | 240 |
| Navarra | 654,214 | 6.6 (5.1; 8.5) | 528 | 1.22 (0.95; 1.58) | 776 (400; 1,494) | 29 | 1.80 (1.39; 2.32) | 248 |
| País Vasco | 2,207,776 | 3.6 (2.9; 4.7) | 1555 | 1.96 (1.50; 2.43) | 1,866 (838; 3,388) | 32 | 2.35 (1.80; 2.91) | 311 |
| La Rioja | 316,798 | 3.7 (2.8; 4.9) | 367 | 3.13 (2.36; 4.14) | 386 (59; 849) | 21 | 3.29 (2.49; 4.35) | 19 |

**Table 2.** Accumulated COVID-19 deaths and MoMo Excess Deaths (*ED*) for Spain and its 19 regions. Values from 23 June to 29 November 2020. The percentages for this period were estimated as differences between the accumulated percentages on 29 November and on 22 June.

| Region | Percentage of infections<sup>14</sup> | Official data |  | MoMo data |  |  | *ED* minus COVID-19 deaths |
| --- | --- | --- | --- | --- | --- | --- | --- |
|  | % (CI 95%) | COVID-19 deaths | CFR (CI 95%) | *ED* (CI 99%). | Days with lower limit >0 | CFR (CI 95%) |  |
| Spain | 4.9 (4.6; 5.2) | 17439 | 0.76 (0.71; 0.81) | 23,612 (11,356; 39,690) | 104 | 1.02 (0.97; 1.09) | 6173 |
| Andalucía | 4.1 (3.5; 4.7) | 3007 | 0.87 (0.76; 1.02) | 5,814 (2,173; 10,738) | 77 | 1.69 (1.47; 1.97) | 2807 |
| Aragón | 7.1 (5.9; 8.4) | 1416 | 1.51 (1.28; 1.82) | 1,666 (421; 3,263) | 59 | 1.78 (1.50; 2.14) | 250 |
| Asturias | 3.7 (2.8; 5.1) | 709 | 1.87 (1.36; 2.48) | 1,198 (320; 2,933) | 31 | 3.17 (2.30; 4.18) | 489 |
| Islas Baleares | 4.7 (3.4; 6.3) | 218 | 0.40 (0.30; 0.56) | 470 (18; 1,666) | 6 | 0.87 (0.65; 1.20) | 252 |
| Canarias | 1.4 (1.0; 2.0) | 178 | 0.59 (0.41; 0.83) | 592 (31; 2,227) | 9 | 1.96 (1.37; 2.75) | 414 |
| Cantabria | 2.7 (2.0; 3.6) | 109 | 0.70 (0.52; 0.94) | 287 (6; 1,127) | 4 | 1.83 (1.37; 2.47) | 178 |
| Castilla - La Mancha | 7.2 (6.6; 8.0) | 1020 | 0.70 (0.63; 0.76) | 1,601 (214; 3,748) | 36 | 1.09 (0.98; 1.19) | 581 |
| Castilla y León | 4.6 (4.1; 5.1) | 1843 | 1.67 (1.51; 1.87) | 2,053 (481; 4,397) | 47 | 1.86 (1.68; 2.09) | 210 |
| Cataluña | 6.1 (5.1; 7.1) | 1914 | 0.41 (0.35; 0.49) | 3,677 (777; 8,125) | 37 | 0.79 (0.67; 0.94) | 1763 |
| Ceuta | 8.9 (6.2; 12.2) | 51 | 0.68 (0.49; 0.97) | 144 (6; 276) | 4 | 1.90 (1.39; 2.73) | 93 |
| C. Valenciana | 3.4 (2.7; 4.2) | 970 | 0.57 (0.46; 0.72) | 2,235 (197; 5,772) | 29 | 1.31 (1.06; 1.65) | 1265 |
| Extremadura | 4.3 (3.7; 5.0) | 443 | 0.97 (0.83; 1.12) | 965 (72; 2,480) | 16 | 2.10 (1.81; 2.44) | 522 |
| Galicia | 2.4 (1.8; 3.2) | 600 | 0.93 (0.70; 1.23) | 1,148 (64; 3,972) | 10 | 1.77 (1.33; 2.36) | 548 |
| Madrid | 8.2 (7.3; 9.1) | 2757 | 0.51 (0.46; 0.57) | 2,930 (793; 6,373) | 61 | 0.54 (0.48; 0.60) | 173 |
| Melilla | 8.4 (6.1; 11.3) | 39 | 0.54 (0.40; 0.74) | 135 (6; 260) | 5 | 1.86 (1.38; 2.56) | 96 |
| Murcia | 4.0 (2.8; 5.4) | 478 | 0.80 (0.60; 1.14) | 785 (58; 2,212) | 13 | 1.31 (0.97; 1.88) | 307 |
| Navarra | 7.9 (6.8; 9.1) | 361 | 0.70 (0.61; 0.81) | 472 (24; 1,474) | 7 | 0.91 (0.79; 1.06) | 111 |
| País Vasco | 4.4 (3.6; 5.3) | 1038 | 1.07 (0.89; 1.31) | 1,311 (174; 3,494) | 25 | 1.35 (1.12; 1.65) | 273 |
| La Rioja | 5.2 (3.4; 7.6) | 183 | 1.12 (0.76; 1.71) | 341 (24; 956) | 12 | 2.07 (1.42; 3.17) | 158 |

**Figure 2.**
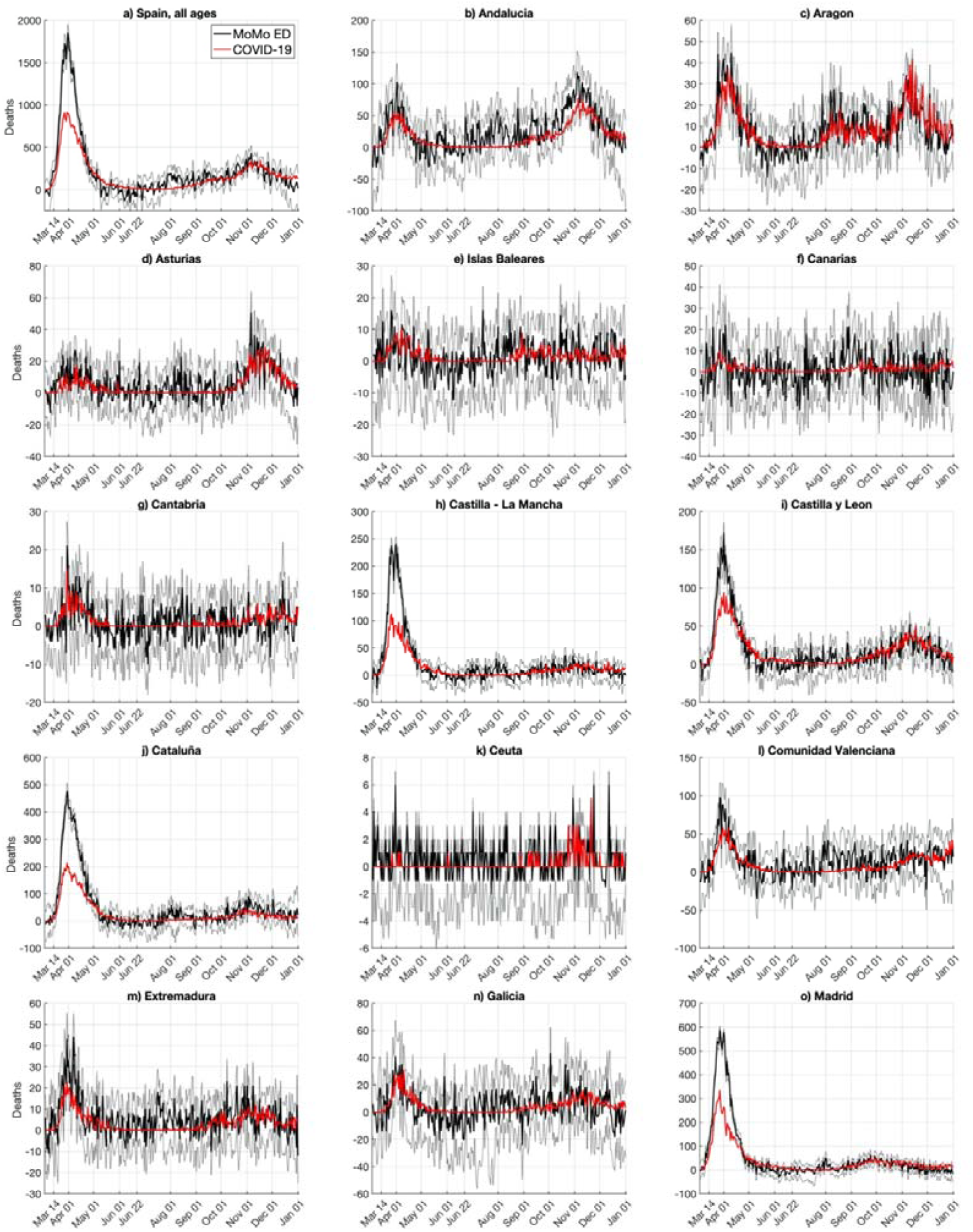
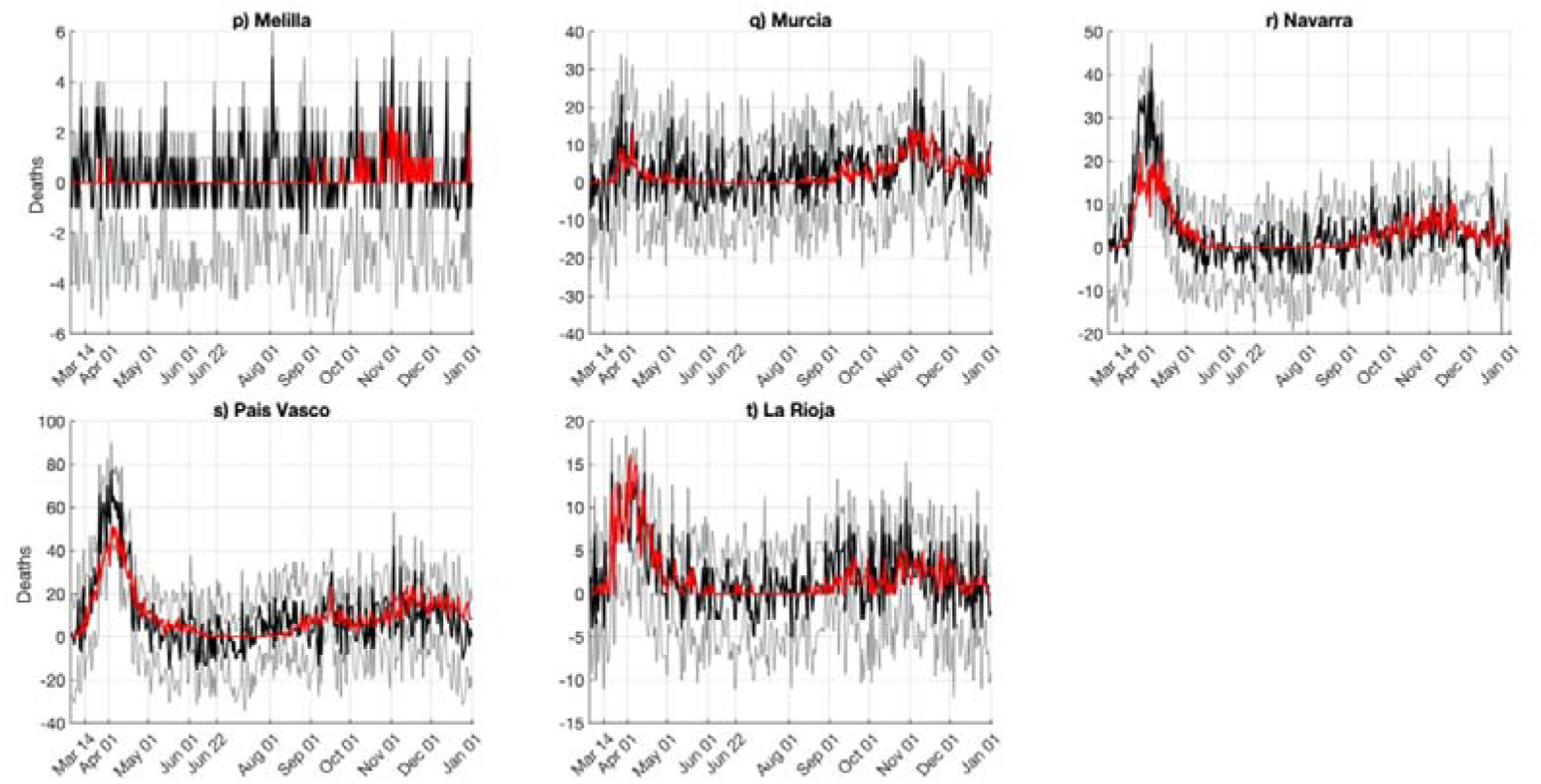
COVID-19 deaths and MoMo Excess Deaths (*ED*). a) Spain; b-t) Spanish regions. Red lines are official COVID-19 deaths, solid black lines are MoMo deaths, and thin black lines delimit MoMo death error bands (99% confidence interval).

### 3.2. Infections estimated from COVID-19 deaths

To illustrate the delay between official daily infections data and REMEDID estimated daily infections, we applied REMEDID from COVID-19 deaths assuming CFR= 100%. Figure 3 shows the current *IO_21_* and the infections associated with COVID-19 deaths for the first wave. The latter in Spain reached a maximum on 13 March 2020 (Table 3), the day before the national government decreed a state of emergency and national lockdown. Thus, the adopted measures had an immediate effect, which was observed in the official data *IO_21_* 7 days later (20 March). This delay is similar to the incubation period (mean 5.78 days^2^), which could be explained because official infections were reported when symptoms appeared. This delay reached 16 days when we compared with earlier version *IO_20_* (not shown), which highlights the usefulness of the methodology to reinterpret official data from very early stages of the pandemic. On the other hand, the maximum number of deaths was reached on 1 April, which was 19 days after the inferred infection maximum, bringing this delay close to the 20 days expected between infection and death (Figures 1 and 3).

**Figure 3.**
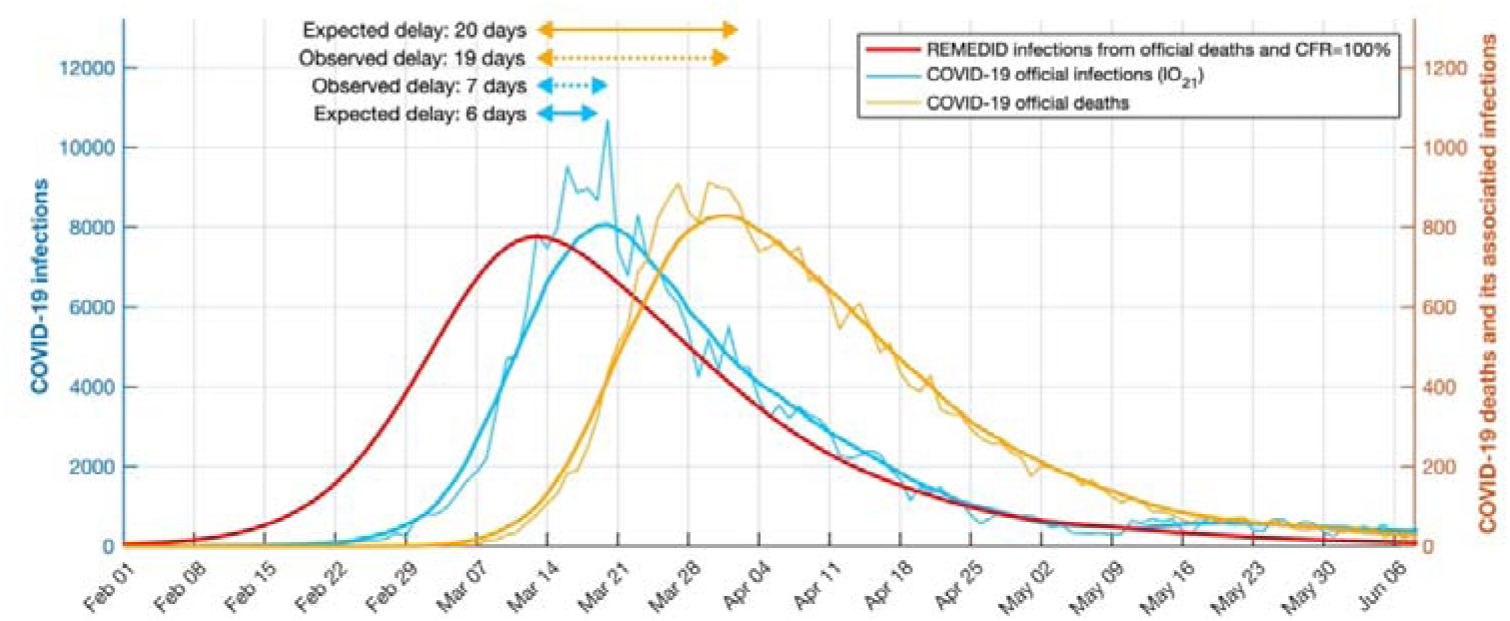
Official COVID-19 infections and deaths, and estimated infections with case fatality ratio (CFR) of 100% in Spain during the first wave. Left y-axis: COVID-19 daily infections IO_21_ (blue curve). Right y-axis: COVID-19 deaths (orange curve) and its REMEDID-estimated infections with CFR=100% (red curve). All curves are for Spain. Thin blue and orange curves are daily data, and thick curves are smoothed by 14- day running mean. Arrows show delays between the maximum of inferred infections and maxima from COVID-19 deaths (orange arrows) and COVID-19 infections (blue arrows). Solid arrows are expected delays, dotted while arrows are observed delays.

We applied REMEDID to the official COVID-19 death with the corresponding estimated CFRs (see section 2.2) to obtain the time series of estimated daily infections, hereafter referred to as *IR_O_*. Figure 4 shows *IR_O_* and the accumulated infections for Spain and its 19 regions. Note that in Spain, *IR_O_* are amplified versions of inferred infections in Figure 3. In Spain, the first infection, according to *IR_O_*, is on 8 January 2020 (Table 3), 43 days before the first infection was officially reported on 20 February 2020 according to *IO_20_*. By contrast, *IO_21_* places the first infection on 1 January 2020. Spain reached the maximum number of *IR_O_* on 13 March, a day before the state of emergency and lockdown were enforced (Table 4). On 14 March, *IO_20_* = 1,832, and *IO_21_* = 7,478; however, *IR_O_* = 63,727 (CI 95%: 60,050-67,403), 35 and 9 times *IO_20_* and *IO_21_*, respectively (Table 5). This implies that on that day, *IO_20_* and *IO_21_* only reported 2.9% (CI 95%: 2.7%-3.1%) and 11.7% (CI 95%: 11.1%-12.5%) of new infections, respectively. Although detection of infections clearly improved from *IO_20_* to *IO_21_*, almost 90% of the infections are still not documented in the peak of the first wave. The situation is similar for the accumulated infections before 22 June 2020, as reported by the National Seroprevalence Study^4, 5^.

**Table 4.** Date of the most prominent relative maxima, for Spain and the 19 regions, of the REMEDID estimated daily infections from COVID-19 deaths (*IR_O_*) and from MoMo Excess Deaths (*ED*) (*IR_M_*), and official COVID-19 daily infections (*IO_21_*). Maxima of *IO_21_* were estimated from the 14-day running mean time series.

|  | Relative Maxima in 2020 |  |  |
| --- | --- | --- | --- |
| | $IR_O$ | $IR_M$ | $IO_{2I}$ |
| Spain | 13 March, 22 October | 13 March, 18 July, 30 Aug, 19 Oct | 20 March, 16-17 Sept, 26 Oct |
| Andalucía | 15 March, 21 October | 12 March, 19 October | 20 March, 26 October |
| Aragón | 19 March, 27 July, 24 Oct | 16 March, 25 July, 21 Oct | 23 March, 2 Aug, 26 Oct |
| Asturias | 25-26 March, 2-3 Nov | 12-14 March, 22-23 March, 25 July, 24 Oct | 20 March, 9 Nov |
| Islas Baleares | 17 March, 16 Aug, 5 Sept, 20-21 Oct | 12 March, 30-31 July, 3 Oct | 20 March, 17 Aug, 8 Nov, 20-21 Nov |
| Canarias | 11 March, 26-27 Aug | 14 March, 12 Aug | 19 March, 29 Aug |
| Cantabria | 17 March, 2 Sept, 26 Oct, 16 Nov | 17 March | 22 March, 4 Sept, 8 Nov |
| Castilla - La Mancha | 12 March, 19 Oct | 12 March, 18 July, 15 Oct | 20 March, 17 Sept, 29 Oct |
| Castilla y León | 15 March, 25 Oct | 14 March, 21-22 Oct | 21 March, 26 Oct |
| Cataluña | 14 March, 12 Oct | 15 March, 22 July, 19 Oct | 20 March, 26 Oct |
| Ceuta | 17-19 March, 16-17 May, 25-26 Aug, 15-16 Oct | 22-24 July, 24-26 Aug, 14-15 Oct | 23 March, 29 Oct |
| C. Valenciana | 15 March, 1 Sept, 31 Oct | 13 March, 18 July, 24 Nov | 19 March, 24 Aug, 2 Nov |
| Extremadura | 14 March, 12-13 Sept, 19 Oct | 14 March, 3 July, 13 Oct | 22 March, 18 Sept, 17 Oct |
| Galicia | 18 March, 28-29 Aug, 25 Oct | 17 March, 8-9 Oct | 22 March, 29 Aug, 27 Oct |
| Madrid | 11 March, 9 Sept | 11 March, 18 July, 8 Sept | 28 March, 16 Sept |
| Melilla | 11 March, 20 Aug, 16 Oct | 9-10 March, 17-18 July, 15 Oct | 17 March, 8 Sept, 22 Oct |
| Murcia | 14-15 March, 28-29 Aug, 19-20 Oct | 11-12 March, 17 April, 18-19 Aug, 17-18 Oct | 19 March, 18 Sept, 30 Oct |
| Navarra | 16 March, 21 Oct | 14 March, 23 Sept, 17 Oct | 19 March, 17 Sept, 20 Oct |
| País Vasco | 18 March, 31 Aug, 30-31<br>Oct | 15-16 March, 30 Aug, 16 Oct | 20 March, 30 Aug, 1 Nov |
| La Rioja | 18 March, 3-4 Sept, 22-23<br>Oct | 13-14 March, 21-22 Aug, 10-11<br>Oct | 21 March, 14 Sept, 26 Oct |

**Table 5.** REMEDID estimated daily infections from COVID-19 deaths (*IR_O_*) and from MoMo Excess Deaths (*ED*) (*IR_M_*) on 14 March, and for official COVID-19 daily infections released on June 2020 (*IO_20_*) and released on February 2021 (*IO_21_*). Infections are inferred from COVID-19 deaths and MoMo ED with their corresponding CFR and 95% confidence interval detailed in Table 1. Percentages are official COVID-19 infections as proportions of the estimated infections.

|  | Daily infections on 14 March 2020 |  |  |  | Percentage of detected daily infections (%) |  |  |  |
| --- | --- | --- | --- | --- | --- | --- | --- | --- |
| | $IR_D$ | $IR_M$ | Official COVID-19 | | $IO_{20}$ | | $IO_{21}$ | |
| | (CI 95%) | (CI 95%) | $IO_{20}$ | $IO_{21}$ | $IR_D$ | $IR_M$ | $IR_D$ | $IR_M$ |
|  |  |  |  |  | (CI 95%) | (CI 95%) | (CI 95%) | (CI 95%) |
| Spain | 63,727<br>(60,050; 67,403) | 77855 (73364;<br>82347) | 1832 | 7478 | 2.9 (2.7; 3.1) | 2.4 (2.2;<br>2.5) | 11.7 (11.1;<br>12.5) | 9.6 (9.1;<br>10.2) |
| Andalucía | 7,144<br>(6,192; 8,573) | 6788 (5883; 8145) | 168 | 396 | 2.4 (2.0; 2.7) | 2.5 (2.1;<br>2.9) | 5.5 (4.6;<br>6.4) | 5.8 (4.9;<br>6.7) |
| Aragón | 1,599<br>(1,232; 2,065) | 1804 (1391; 2330) | 26 | 107 | 1.6 (1.3; 2.1) | 1.4 (1.1;<br>1.9) | 6.7 (5.2;<br>8.7) | 5.9 (4.6;<br>7.7) |
| Asturias | 358<br>(245; 490) | 346 (237; 473) | 45 | 81 | 12.6 (9.2; 18.4) | 13 (9.5; 19) | 22.6 (16.5;<br>33.1) | 23.4 (17.1;<br>34.2) |
| Islas<br>Balears | 412<br>(235; 706) | 307 (175; 526) | 14 | 69 | 3.4 (2.0; 6.0) | 4.6 (2.7; 8) | 16.7 (9.8;<br>29.4) | 22.5 (13.1;<br>39.4) |
| Canarias | 1,385 (903;<br>2,108) | 1086 (708; 1653) | 19 | 96 | 1.4 (0.9; 2.1) | 1.7 (1.1;<br>2.7) | 6.9 (4.6;<br>10.6) | 8.8 (5.8;<br>13.6) |
| Cantabria | 597 (403; 871) | 501 (338; 730) | 20 | 59 | 3.4 (2.3; 5.0) | 4 (2.7; 5.9) | 9.9 (6.8;<br>14.6) | 11.8 (8.1;<br>17.5) |
| Castilla - La<br>Mancha | 5,303 (4,309;<br>6,408) | 6458 (5247; 7804) | 112 | 545 | 2.1 (1.7; 2.6) | 1.7 (1.4;<br>2.1) | 10.3 (8.5;<br>12.6) | 8.4 (7;<br>10.4) |
| Castilla y<br>León | 4,688 (4,087;<br>5,410) | 5871 (5119; 6775) | 54 | 474 | 1.2 (1.0; 1.3) | 0.9 (0.8;<br>1.1) | 10.1 (8.8;<br>11.6) | 8.1 (7; 9.3) |
| Cataluña | 11,795 (9,996;<br>14,194) | 13854 (11740;<br>16671) | 184 | 1482 | 1.6 (1.3; 1.8) | 1.3 (1.1;<br>1.6) | 12.6 (10.4;<br>14.8) | 10.7 (8.9;<br>12.6) |
| Ceuta | 23 (10; 59) | 9 (4; 23) | 1 | 3 | 4.3 (1.7; 10.0) | 11.1 (4.3;<br>25) | 13.0 (5.1;<br>30) | 33.3 (13;<br>75) |
| C.<br>Valenciana | 3,476 (2,752;<br>4,490) | 3555 (2814; 4592) | 279 | 463 | 8.0 (6.2; 10.1) | 7.8 (6.1;<br>9.9) | 13.3 (10.3;<br>16.8) | 13 (10.1;<br>16.5) |
| Extremadura | 981 (696; 1,361) | 883 (627; 1225) | 29 | 71 | 3.0 (2.1; 4.2) | 3.3 (2.4;<br>4.6) | 7.2 (5.2;<br>10.2) | 8 (5.8;<br>11.3) |
| Galicia | 1,476 (1,165;<br>1,865) | 1443 (1139; 1823) | 80 | 244 | 5.4 (4.3; 6.9) | 5.5 (4.4; 7) | 16.5 (13.1;<br>20.9) | 16.9 (13.4;<br>21.4) |
| Madrid | 19,565 (17,224;<br>22,241) | 24315 (21405;<br>27640) | 604 | 2622 | 3.1 (2.7; 3.5) | 2.5 (2.2;<br>2.8) | 13.4 (11.8;<br>15.2) | 10.8 (9.5;<br>12.2) |
| Melilla | 139 (94; 208) | 46 (31; 68) | 5 | 3 | 3.6 (2.4; 5.3) | 10.9 (7.4;<br>16.1) | 2.2 (1.4;<br>3.2) | 6.5 (4.4;<br>9.7) |
| Murcia | 867 (542; 1,464) | 446 (279; 752) | 24 | 84 | 2.8 (1.6; 4.4) | 5.4 (3.2;<br>8.6) | 9.7 (5.7;<br>15.5) | 18.8 (11.2;<br>30.1) |
| Navarra | 1,165 (900;<br>1,500) | 1447 (1118; 1863) | 37 | 121 | 3.2 (2.5; 4.1) | 2.6 (2; 3.3) | 10.4 (8.1;<br>13.4) | 8.4 (6.5;<br>10.8) |
| País Vasco | 1,965 (1,583;<br>2,566) | 2269 (1828; 2962) | 109 | 421 | 5.5 (4.2; 6.9) | 4.8 (3.7; 6) | 21.4 (16.4;<br>26.6) | 18.6 (14.2;<br>23) |
| La Rioja | 314 (238; 416) | 266 (201; 352) | 22 | 137 | 7.0 (5.3; 9.2) | 8.3 (6.3;<br>10.9) | 43.6 (32.9;<br>57.6) | 51.5 (38.9;<br>68.2) |

In almost all regions, *IO_20_* showed a delay of 1 month or more between the first infection and *IR_O_* (Table 3). No delay in *IO_21_* occurred in Islas Baleares, Castilla-León, and Galicia, while in three regions (Cataluña, Madrid, and La Rioja), the first case occurred earlier than the first case of *IR_O_*. However, 6 regions had delays of 15 days and other 6 regions had delays of 1 month. According to *IR_O_*, all regions except Ceuta and Melilla had some infections in January, but in *IO_21_* only 6 regions had infections in that month. In all scenarios, the first infections were in Madrid and Cataluña.

During the first wave, according to *IR_O_* most of the regions had maximum daily infections around 14 March. In Madrid, the maximum was reached on 11 March, coinciding with the educational centres closing and an official warning by the regional government (Table 4). Asturias was the last region to reach peak infections (25-26 March). The maximum percentage of documented cases (12.6%,CI 95%: 9.2%-18.4%) occurred in Asturias on 14 March, but in the other regions, only between 1.2 to 8 % of the infections were documented (Table 5).

Figure 4 shows how the *IO_21_* and *IR_O_* curves of Spain and the 19 different regions fluctuated following the same pattern until the middle of June 2020, but thereafter, they showed different patterns. This reflects the fact that the Spanish government had decreed the control measures for the whole nation until June, but thereafter, each regional government implemented its own control measures. For example, some regions (e.g., Aragón, Islas Baleares, Cantabria, Comunidad Valenciana, Extremadura, Galicia, Murcia, País Vasco, and La Rioja) had two peaks, but others had only one. An apparent maximum on 22 June in Islas Baleares is an artifact produced by the interpolation for transition from the two CFRs. Although beyond the scope of this work, it would be very interesting to investigate the effects of the different control measures implemented on the corresponding *IR_O_* for the 19 regions.

The Spanish COVID-19 second wave reached a maximum of daily infections on 22 October from *IR_O_* and on 26 October from *IO_21_*. The delay of 4 days is similar to the mean incubation period (5.78 days^2^). The estimated number of new infections is still larger than the documented cases, but the shapes of the two curves are more similar in the second wave than in the first wave (Figure 1a). The same is true for the 19 regions, most of which had the largest peak around 22-26 October, with the exceptions of Canarias and Madrid, which reached maxima in late August and early September, respectively.

**Table 3.** Date of first infection for REMEDID estimated daily infections from COVID- 19 deaths (*IR_O_*) and from MoMo Excess Deaths (*ED*) (*IR_M_*), and for official COVID-19 daily infections released on June 2020 (*IO_20_*) and on February 2021 (*IO_21_*). All dates refer to year 2020.

|  | First infection |  |  |  |
| --- | --- | --- | --- | --- |
| | $IR_O$ (CI 95%) | $IR_M$ (CI 95%) | Official COVID-19 data | |
| | | | $IO_{20}$ | $IO_{21}$ |
| Spain | 8 Jan (8-8 Jan) | 9 Jan (9-10 Jan) | 20 Feb | 1 Jan |
| Andalucía | 18 Jan (17-18 Jan) | 12 Jan (12-13 Jan) | 26 Feb | 2 Feb |
| Aragón | 21 Jan (20-22 Jan) | 8 Jan (8 Jan-8 Jan) | 4 March | 11 Feb |
| Asturias | 28 Jan (26-29 Jan) | 20 Jan (18 Jan-21 Jan) | 29 Feb | 23 Feb |
| Islas Baleares | 27 Jan (24-29 Jan) | 14 Jan (11 Jan-17 Jan) | 20 Feb | 29 Jan |
| Canarias | 22 Jan (20-23 Jan) | 8 Jan (8 Jan-8 Jan) | 20 Feb | 29 Jan |
| Cantabria | 28 Jan (26-30 Jan) | 11 Jan (9 Jan-13 Jan) | 29 Feb | 23 Feb |
| Castilla - La Mancha | 17 Jan (16-17 Jan) | 14 Jan (13 Jan-14 Feb) | 29 Feb | 24 Feb |
| Castilla y León | 19 Jan (19-20 Jan) | 17 Jan (16-17 Jan) | 28 Feb | 20 Jan |
| Cataluña | 16 Jan (16-17 Jan) | 10 Jan (10 Jan-11 Feb) | 25 Feb | 3 Jan |
| Ceuta | 16 Feb (11-21 Feb) | 21 Jan (16 Jan-26 Jan) | 14 March | 3 March |
| C. Valenciana | 9 Jan (8-11 Jan) | 12 Jan (11 Jan-13 Jan) | 25 Feb | 1 Feb |
| Extremadura | 25 Jan (24-26 Jan) | 12 Jan (10 Jan-13 Jan) | 29 Feb | 8 Feb |
| Galicia | 25 Jan (24-26 Jan) | 14 Jan (13 Jan-15 Jan) | 3 March | 1 Feb |
| Madrid | 11 Jan (10-11 Jan) | 9 Jan (8 Jan-9 Jan) | 25 Feb | 1 Jan |
| Melilla | 2 Feb (31 Jan-4 Feb) | 17 Jan (15 Jan-19 Jan) | 12 March | 5 March |
| Murcia | 28 Jan (26-29 Jan) | 13 Jan (10 Jan-15 Jan) | 7 March | 22 Feb |
| Navarra | 24 Jan (23-25 Jan) | 8 Jan (8 Jan-8 Jan) | 29 Feb | 10 Feb |
| País Vasco | 18 Jan (17-18 Jan) | 9 Jan (9 Jan-11 Feb) | 28 Feb | 12 Feb |
| La Rioja | 27 Jan (25-28 Jan) | 14 Jan (12 Jan-15 Jan) | 1 March | 5 Jan |

**Figure 4.**
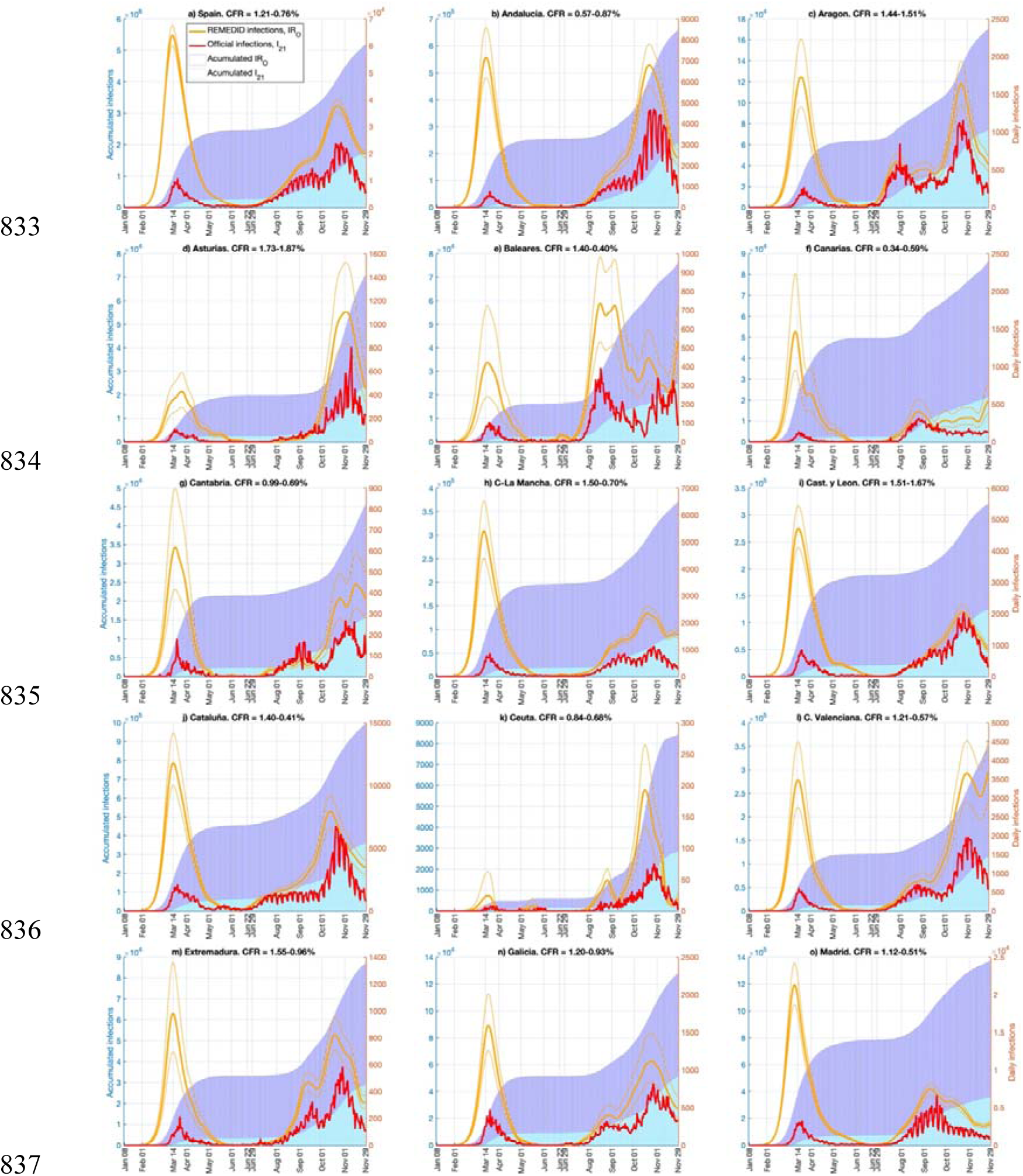
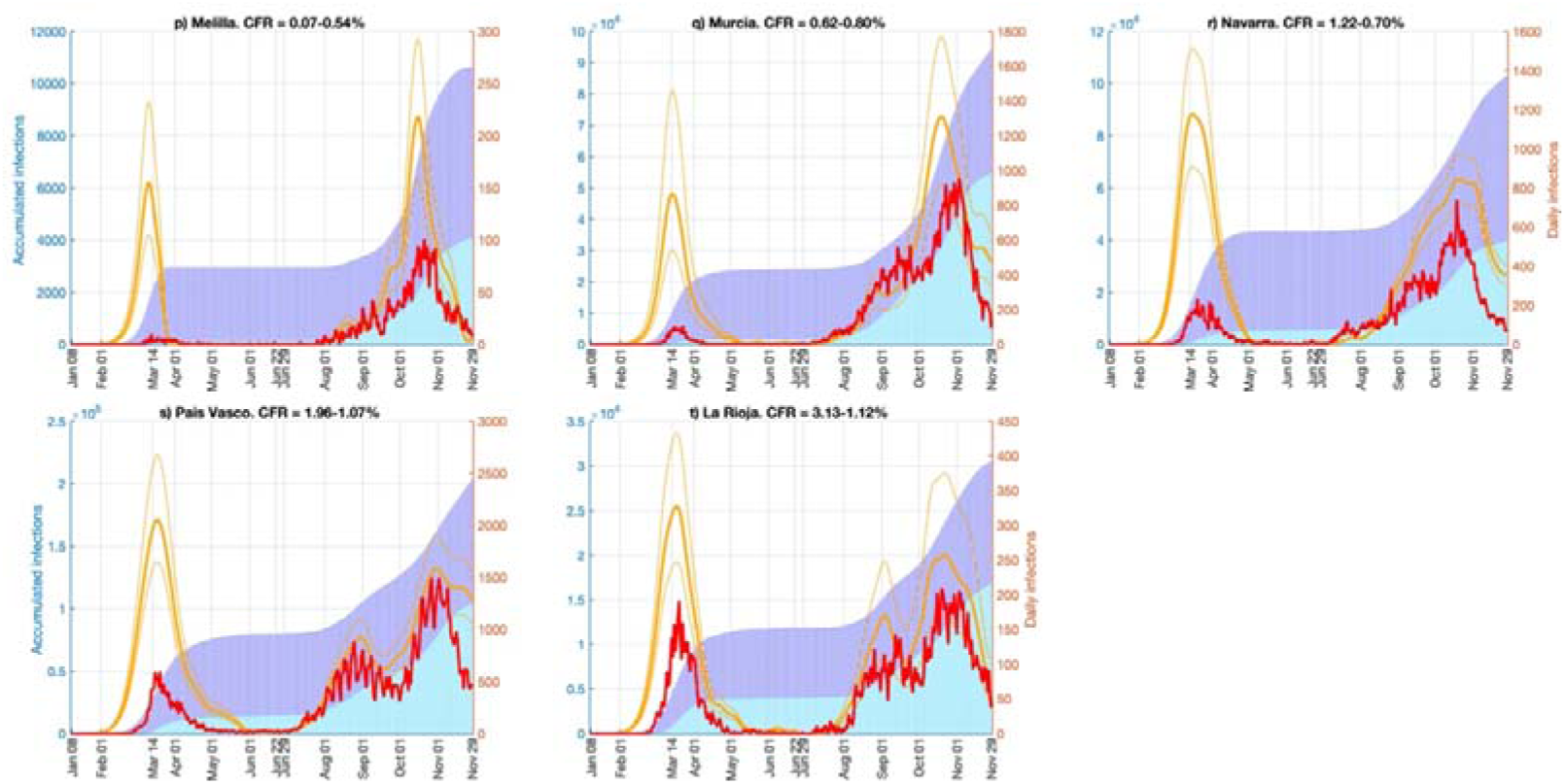
Daily and accumulated infections for official COVID-19 daily infections (*IO_21_*), and daily infections estimated from COVID-19 deaths (*IR_O_*). Lines are daily infections and refer to the y-axis on the right; bars are accumulated infections and refer to the y-axis on the left. Red lines and cyan bars are official COVID-19 data; orange lines and blue bars are inferred infections with case fatality ratio (CFR) in Table 1. Thin orange lines correspond to the CFR confidence interval.

### 3.3. Infections from MoMo Excess Deaths

Assuming that MoMo *ED* accounts for both recorded and non-recorded COVID-19 deaths, negative deaths are meaningless, and they were set to zero. Then, the associated daily infections can be estimated, as in Section 3.2, with a CFR of 100% from MoMo *ED* for Spain (Figure 5). Note two main differences between this time series and that estimated from official COVID-19 deaths: (1) MoMo data present an error band that was inherited by the estimated infections; (2) MoMo *ED* estimated infections reached a maximum of 1,443 (CI 99%: 1,329-1,547), doubling the 776 inferred daily infections from official COVID-19 deaths in Figure 3. This is because maximum MoMo *ED* was 1,584 (CI 99%: 1,468-1,686%) and maximum COVID-19 official deaths was 828, both estimated from the 14-day running mean time series. The maximum of inferred infections was reached on 13 March, just one day prior to the state of emergency and lockdown. The expected and observed delays with respect to official infections and MoMo *ED* were similar to those observed for estimated infections from official COVID-19 deaths. Error bounds of the estimated infections in Figure 5 were computed from the MoMo *ED* error bounds. However, it should be highlighted that the combination of the error bounds from MoMo *ED* and the estimated CFRs might lead to unrealistic error estimates. To avoid this, the error estimates in Figure 6 were estimated from the MoMo *ED* time series (no error bounds) and the error bounds of the estimated CFRs.

**Figure 5.**
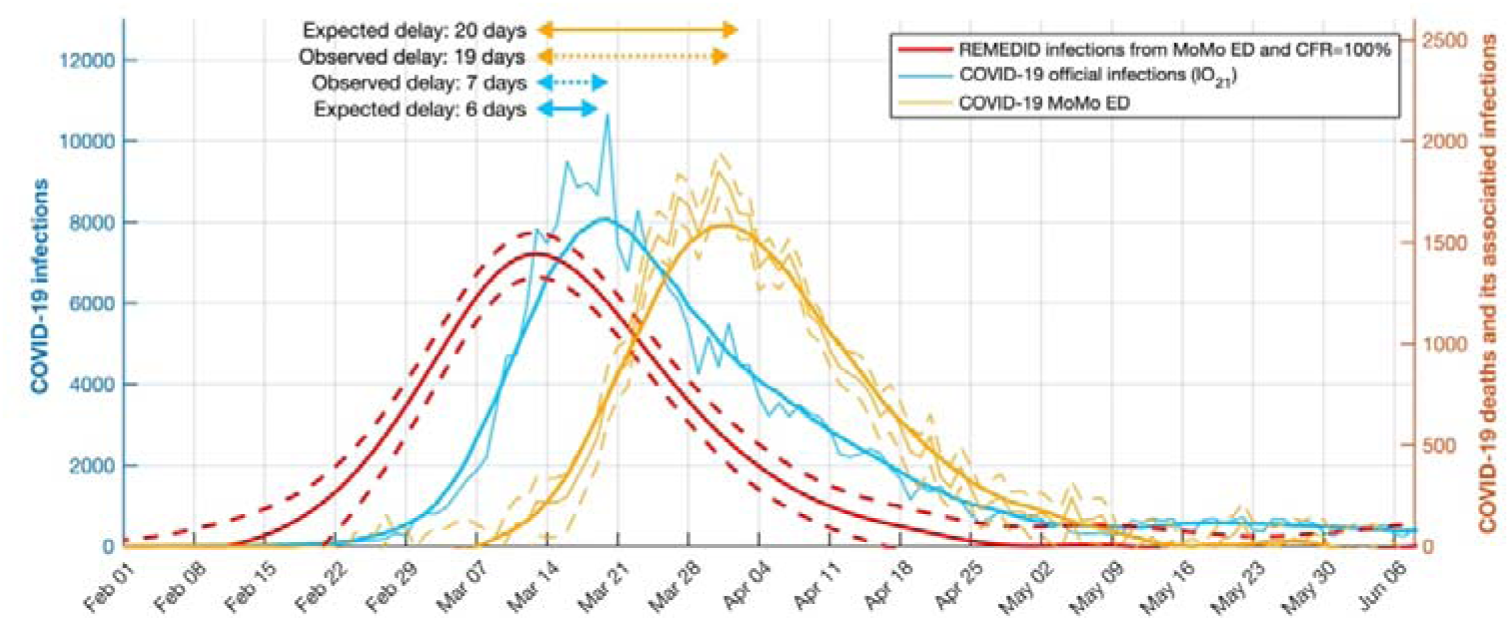
Official COVID-19 infections, MoMo Excess Deaths (*ED*), and estimated infections with case fatality ratio (CFR) of 100% in Spain during the first wave. Left y- axis: COVID-19 daily infections *IO_21_* (blue curve). Right y-axis: MoMo *ED* (orange curve) and its REMEDID-estimated infections with CFR=100% (red curve). All curves are for Spain. Thin blue and orange curves are daily data, and thick curves are smoothed by 14-days running mean. Dashed curves represent the error estimate of MoMo *ED* (orange) and inferred infections (red). Arrows show delays between the maximum of inferred infections and maxima from MoMo *ED* (orange arrows) and COVID-19 infections (blue arrows). Solid arrows are expected delays, dotted while arrows are observed delays.

The REMEDID was applied to the MoMo *ED* with the corresponding CFRs (see section 2.2) to obtain the estimated daily infections, which will be referred hereafter as *IR_M_*. The *IR_M_* were calculated for Spain and its 19 regions and are depicted in Figure 6, as well as the accumulated *IR_M_*. In Spain, the first infection shown by *IR_M_* happened on 9 January, with an error estimate from 9 to 10 January, 41 to 42 days before the first documented infection of *IO_20_* on 20 February 2020 (Table 3). The maximum *IR_M_* was 77,855 (CI 95%: 73,364 - 82,347) reached on 13 March. On 14 March, *IR_M_* showed 14,128 infections more than *IR_O_* (Table 5). Notice that the CFR used with MoMo *ED* data was larger than the one used with official COVID-19 deaths data, which makes this difference even more remarkable, because the larger the CFR the lower the estimated infections. Therefore, if the true CFRs, which are unknown, were used in both cases, *IR_M_* would double *IR_O_* on 14 March, as happened when a CFR of 100% was used (Figures 3 and 5). Notice that with the CFRs used, the *IR_M_* and *IR_O_* resulted in the same accumulated infections on 22 June and 29 November, matching the results of the seroprevalence study. Nevertheless, *IR_M_* showed 42 times more cases than *IO_20_* and 10 times more than *IO_21_* on 14 March, detection of official cases of only 2.4% (2.2%-2.5%) and 9.6% (9.1%-10.2%), respectively.

Table 3 shows the estimated date of first infection for Spain and by region. Note that the first cases of *IR_M_* in Spain were on 9 January and in Aragón, Canarias, and Navarra on 8 January, which is possible because significant excess deaths in a region may not become significant for the whole country. In general, the maxima of daily infections were closer to those on 14 March in *IR_M_* than in *IR_O_*. During the first wave, all regions showed a single maximum, except for Ceuta, Melilla, and Murcia, which showed two maxima (Figure 6). In general, the *IR_M_* time series in all regions were similar during that period. The official data clearly under-detected infections during the first wave. On 14 March, *IR_M_* were comparable to *IR_O_*, overlapping CI in all regions, but not in Spain as a whole (Table 5). During the second wave, there was improved detection of cases with differences among regions.

**Figure 6.**
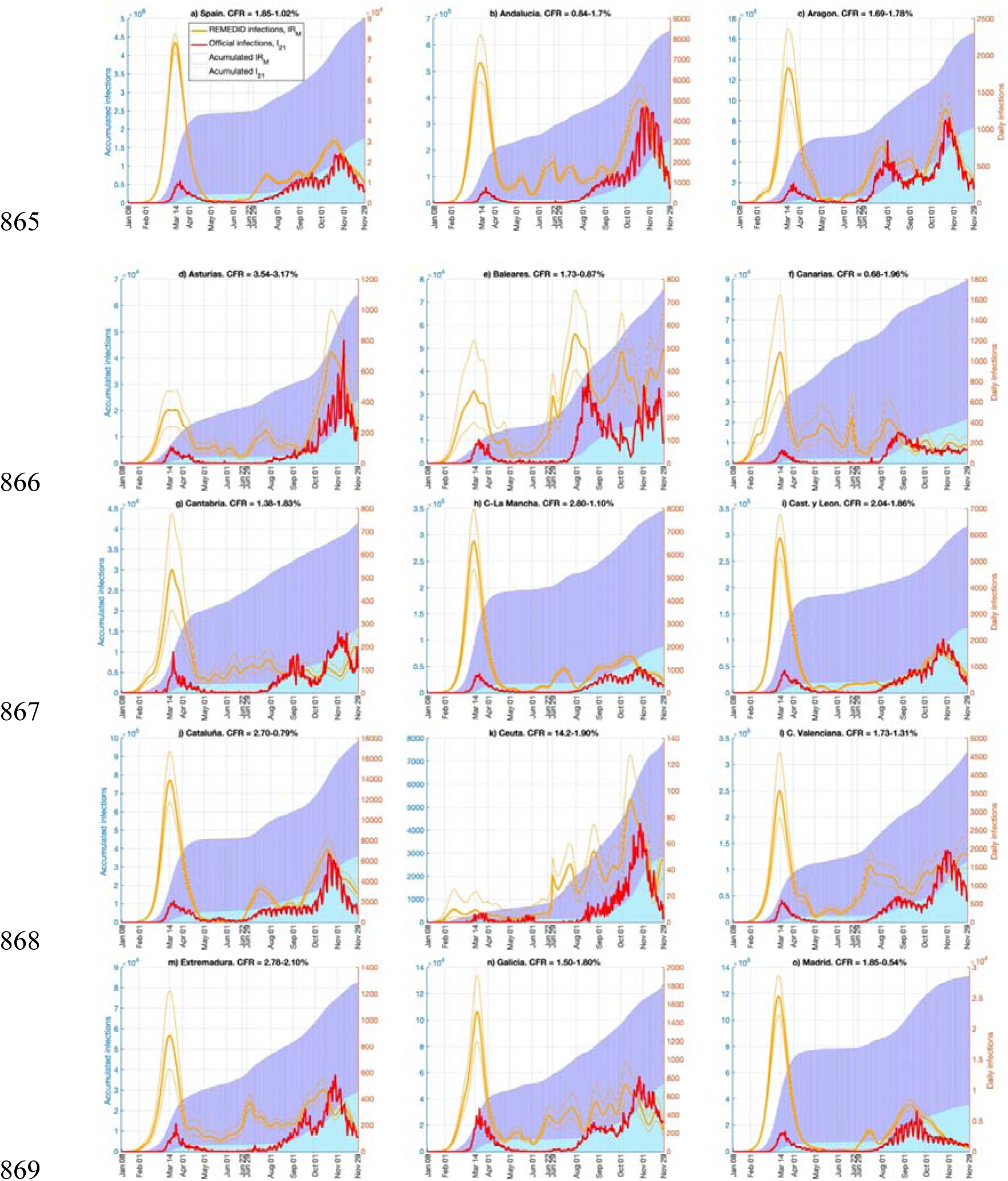
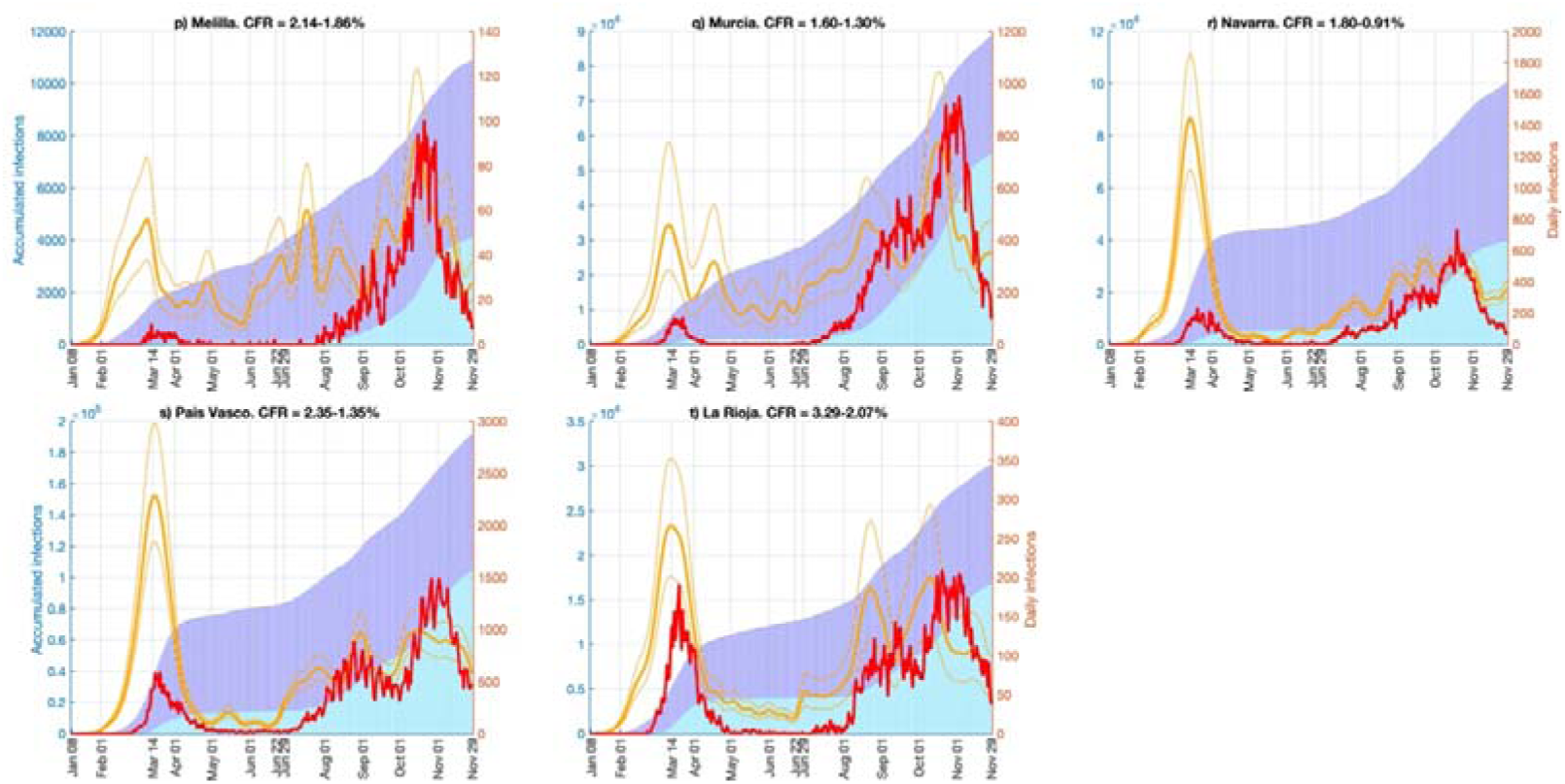
Daily and accumulated infections for official COVID-19 daily infections (*IO_21_*), and daily infections estimated from MoMo Excess Deaths (*ED)* (*IR_M_*). Lines are daily infections and refer to the y-axis on the right; and bars are accumulated infections and refer to the y-axis on the left. Red lines and cyan bars are for official COVID-19 data; and orange lines and blue bars are for inferred infections with case fatality ratio (CFR) in Table 1. Thin orange lines represent the error estimate of inferred infections.

### 4. Discussion

Infection dynamics is key to understand and model the COVID-19 pandemic. Nevertheless, documented infections in Spain (and nearly worldwide) are of limited quality qualitatively and quantitatively. Specifically, reported infections were underestimated and delayed 16 days compared to the estimated date of infection during the first months of the pandemic (according to *IO_20_*) and 6 days in the current version of the data (*IO_21_*). According to the National Seroprevalence Study, only 10.7% of infections were documented in Spain prior to 22 June 2020^4, 5^ and 64.3% from 23 June to 29 November 2020^14^. The huge underreporting during the first wave is mainly due to: (1) the lack of testing for asymptomatic and mildly symptomatic people^3^, which could have been detected with a “test, track and trace” strategy, as done in South Korea, China, and Singapore^17^; (2) deaths outside of the hospitals, with many cases from nursing homes for the elderly, where 6,664 deaths with COVID-19 and 7,082 with similar symptoms, but not officially diagnosed with COVID-19, occurred from January to May 2020^18^. This poor quality of data hinders any options to study the infection dynamics based on official data.

To overcome these difficulties, we propose the REMEDID algorithm, which allows calculation of time series for daily infections from daily death time series, a more reliable source, which can be applied in early stages of any pandemic with only 33 days delay. The REMEDID algorithm needs only three inputs:

(1) PDF of the period from date of infection to date of death. In this study, that was calculated by combining the PDF of the Incubation Period and the Illness Onset to Death period, as estimated by Linton et al.11 for cases in Wuhan, China. The incubation period can be assumed to be similar in China and elsewhere, because it is a characteristic of the virus, but the illness onset to death period could be affected by the type of health care and, thus, by each national health system2 or even ethnicity19. No such information is available for Spain to date, but it would be desirable to use a local distribution of Illness Onset to Death when available for Spain. The calculations could be redone easily by applying the MATLAB code available in the Supplementary Material.
(2) Time series of daily deaths. We used two sources, the official COVID-19 deaths and the MoMo ED, because official COVID-19 deaths have been underestimated, especially during the first wave in the regions of Castilla–La Mancha, Castilla y León, Cataluña, and Madrid, because non-hospital deaths and cases during the early pandemic were somewhat undertested (Table 1 and Figure 2). We expect that undocumented COVID- 19 deaths appeared in the excess of expected deaths from any cause calculated from MoMo15. Interpretation of those results must be made carefully because ED is a statistical variable calculated from what happened in previous years and some expected deaths, such as those produced by traffic and work accidents, may have been reduced due to the national lockdown that began on 14 March 2020 and gradually has been eased. Nonetheless, expected fatalities were very low (traffic and work accidents less than 5 deaths daily) compared to the daily death toll from COVID-19 (300 deaths per day average from March to May). Some non-COVID-19 deaths may have been influenced by other pandemic-related factors22, such as breakdown in medical follow-up (especially chronic conditions and among older patients), delays in attending to the healthcare appointments and/or being attended due to the collapse of the Health System, transplant delay and reduction in donors’ numbers from 7.2 to 1.2 per day23, and the decrease of the interventional cardiology activity between 40% and 81%24. Thus, MoMo ED do not exactly correspond to COVID-19 deaths and must be regarded with caution. A detailed study of the changes in other sources of mortality is recommended. Nevertheless, MoMo ED is a valuable source of data that must be explored considering the enormous bias in official data of daily infections.
(3) CFR estimate. CFR was defined as the ratio of deaths among total infections. Thus, the lower the documented infections, the larger the CFR and under-detection of infections leads to overestimation of the CFR. For example, in Spain on 22 June, official data showed 261,111 accumulated infections and 29,676 deaths, a CFR of 11.37%, which was clearly overestimated because the number of infections was underestimated. The National Seroprevalence Study in Spain with random testing showed that 5.2% of the population (∼47 million) had already been infected4,5. Because CFR also depends on death records, we calculated a CFR of 1.21% (CI 95%: 1.15%- 1.29%) from official COVID-19 deaths and a CFR of 1.85% (CI 95%: 1.74%-1.96%) from MoMo ED. However, the CFR is not constant in time nor in space. Different regions showed different CFRs and the fourth phase of the seroprevalence study showed a general decrease in the CFRs for the period 23 June – 29 November. In Spain, the CFR decreased from 1.21% to 0.76% (CI 95%: 0.71%-0.81%) from official COVID-19 deaths and from 1.85% to 1.02% (CI 95%: 0.97%-1.09%) from MoMo ED. This decline is probably due to the improvement of COVID-19 treatments and experience gained since the beginning of the pandemic25. Similar seroprevalence studies are desirable in other countries, because CFRs from official data are so variable. For example, on 22 June 2020, CFRs of 0.6, 5.2, and 14.5% were reported for Iceland, the world, and Italy, respectively26. In addition, we assumed a constant CFR prior to 22 June and from 23 June to 29 November 2020, although introducing time dependence on the CFR would enhance the infected estimates. A constant CFR does not consider who is affected by saturation of the health systems due to limited human and material resources. However, to estimate a time-variable CFR requires accurate infection records among other variables, which are not currently available. Moreover, CFRs among severe cases (mostly detected) are much greater than those among asymptomatic and mild cases (mostly undetected). Consequently, the CFR calculation has a high degree of uncertainty. Nevertheless, because information on the CFRs is expected to improve, calculations in this study can be updated easily using the MATLAB code for REMEDID algorithm provided as Supplementary Material.

We ran the REMEDID algorithm to provide the estimated time series of infections for Spain and for its 19 regions for the period from 8 January to 29 November 2020. These time series provided valuable information to understand the time evolution of the pandemic. Our main findings for Spain are:

(1) On 14 March, when the national government decreed the state of emergency and national lockdown, estimated infections were between 35 and 42 times larger than officially documented ones at that time, and 9-10 times more than current official data.
(2) The national lockdown had a strong effect on the transmission of the virus, as shown by a rapid slope decrease around day 13 March.
(3) The first infection was better determined from inferred infections than from official records during the first months of the pandemic.

The REMEDID algorithm has strengths and limitations. First, it uses the number of deaths, which are more accurately recorded than infections. Second, it allows elucidation of the date of the infections estimated during the seroprevalence studies, which only determines the total number of infections. Thus, the REMEDID algorithm complements the seroprevalence studies. Third, the estimated daily infections place the probable date of infection more accurately than the official numbers. Note that official infections are delayed by the incubation period (from infection to illness onset).

Therefore, although the maxima of infections theoretically should have coincided with the state of emergency and the national lockdown on 14 March, official infections in Spain were maximum 6 days later. Infections estimated from death data do not show such delay, however. Determination of the actual day of infection is very important for evaluation of the immediate effect of the implemented countermeasures. In Madrid the estimated infections reached their maximum on 11 March when regional government warned the population to stay at home and schools and universities were closed, forcing 1.2 million students to stay at home. Moreover, overall recommendations on disease control and social distancing were given by the Ministry of Health to the general public since de beginning of March (see https://www.mscbs.gob.es/en/profesionales/saludPublica/ccayes/alertasActual/nCov-China/ciudadania.htm).

Four, the REMEDID algorithm can be applied to any region, regardless of population size, but being cautious with small populations. For example, in this study we applied REMEDID methodology to a small city (Ceuta with 84,777 inhabitants), a medium size region (Madrid with more than 6.6 million inhabitants), and a country (Spain with 47 million inhabitants). This versatility allows study of different epidemic dynamics as reflected in the variety of shapes and slopes in the curves depicting death and infection records, that show the heterogeneity of the epidemic at each region with different population size and density, social behaviour and transmission pattern, especially during the second wave, showing unique dynamics that must be addressed individually. Knowledge of the real epidemic dynamics of different population nodes is key to succeed in modelling attempts, because we could calculate the sole effect of group size^27^. This spatially-explicit information, in combination with population size per node and mobility would allow us to use a metapopulation approach in future models^28–30^.

Among the methodology limitations, the most important is that it can only be implemented retrospectively. Thus, these estimates cannot be used to control the pandemic in real time. But considering that different regions or countries are at different stages at the same time, results of this methodology for the first communities could be applied elsewhere. Furthermore, it is useful to enhance models and improve our knowledge of the pandemic dynamics, including the effectiveness of the different measures adopted to flatten the curve and design safe post-lockdown measures. In addition, more realistic and accurate models could be obtained by use of the more realistic daily number of infections provided by REMEDID, which, in turn, would improve the models outcome and enhance comparisons of different lockdown and post- lockdown measures^31^.

We believe that REMEDID methodology applied to daily COVID-19 deaths (if accurately reported) or to MoMo *ED* could be useful to analyse the dynamics of the pandemic retrospectively and more accurately quantify the real daily infections with respect to the official numbers. We only need the CFR and, for greater precision, the PDF of Infections to Death or, alternatively, the PDF of Incubation Period and Illness Onset to Death. This approach could be implemented anywhere, improving our knowledge and understanding of the dynamics of the pandemic and the effectiveness of the confinement measures. This is of high importance to develop successful strategies to face and reduce the effects of future epidemic episodes.

## Supporting information

Supplementary material - REMEDID MATLAB code

## Data Availability

All data used is available in the manuscript or in the references.
The MATLAB code is available in the Supplementary Material.

## Appendix A. Validation test

In order to validate REMEDID, it would be desirable to have an accurate time series of daily infections and deaths. Then, the REMEDID infections inferred from deaths could be compared to the recorded infections. However, such datasets are not available because many infections in any disease or plague are usually undetected. On the other hand, REMEDID-estimated infections cannot be compared to those of dynamical models, which also need accurate data to be tuned. In fact, the main motivation for developing REMEDID has been to provide (more) reliable data to feed dynamical models.

As an alternative of validation, we propose the following experiment as a proof of concept. In the official COVID-19 time series, *IO_21_*, infections are given the date of symptoms onset. Then, REMEDID applied to official deaths with the PDF of illness onset to death, *h(t)*, instead of the PDF of infection to death, *f(t)*, should produce a time series comparable to *IO_21_*. Official accumulated cases and deaths on 22 June 2020 were 261,111 and 29,676, respectively, which gives a CFR of 11.37%. This CFR is overestimated due to an under detection of cases in that period. On the other hand, official accumulated cases and deaths from 23 June to 29 November 2020 were 1,482,293 and 17,439, which gives a CFR of 1.18%. Although closer to reality, this value is still overestimated. In any case, if we use these CFR values to construct the segmented timeseries of CFRs, as in section 2.2, when applying REMEDID the resulting estimated daily infections should be comparable to *IO_21_*. The results are shown in Figure A.1 where the REMEDID estimated daily infections is in very good agreement with the smoothed *IO_21_* (14-day running mean), with a correlation between the two series of 0.98 % (p-value<0.001). Because smoothed time series present a high degree of serial correlation, the correlation and the significance level was estimated according to a specific Monte Carlo analysis based on the randomization of phases in the frequency domain^32^. Notice that (i) application of the 14-day running mean to the *IO_21_* series removes the weekly variability due to the weekend infections misreport; (ii) the agreement of the two series would improve with an improved estimate of the CFR time series. Thus, the good match between the estimated and the original daily infections timeseries proves the theoretical concept that supports REMEDID.

**Figure A.1.**
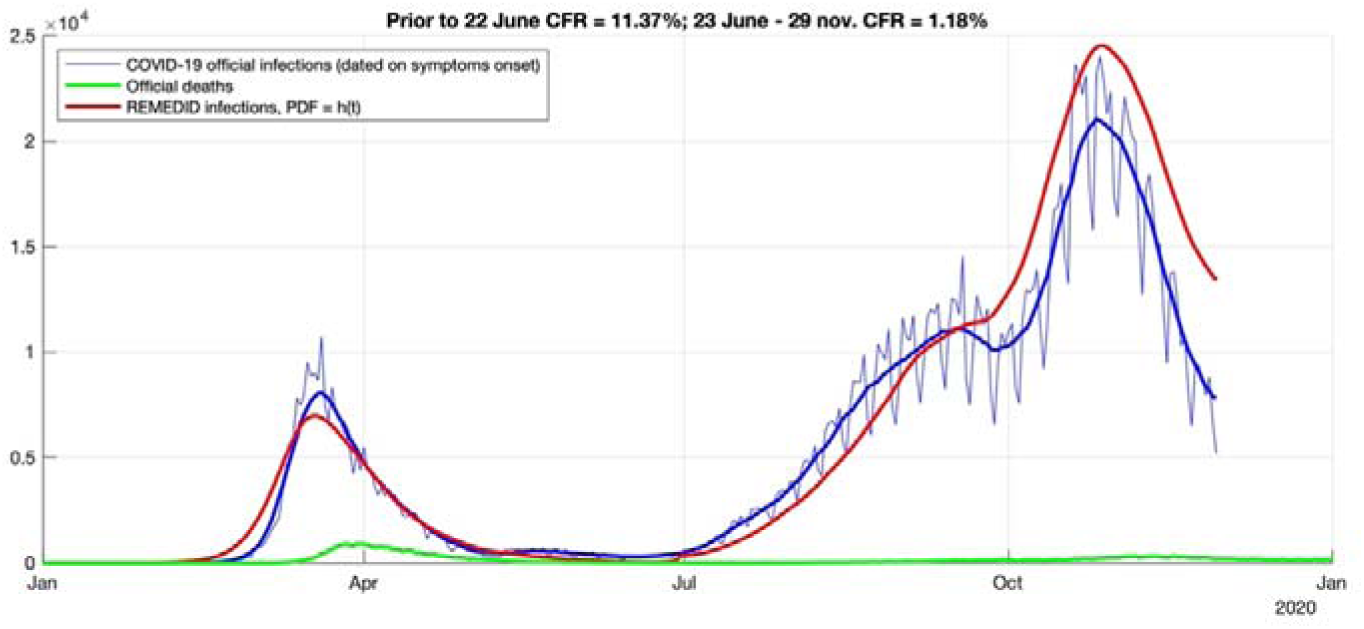
REMEDID validation test. Thin blue line is official infections and thick blue line is its 14-day-running mean; green line is official deaths; and red line is REMEDID applied with the PDF of illness onset to death, *h(t)*.

Note the differences with the results presented in Section 3.2, where the PDF includes the incubation period and a more realistic CFR, as estimated from the National Seroprevalence Study data.

## Notes

### Competing Interest Statement

The authors have declared no competing interest.

### Funding Statement

Funding
This work was supported by the University of Alicante [COVID-19 2020-41.30.6P.0016 to CB] and the Ajuntament de Denia through the Montgo-Denia Research Station Agreement [2020-41.30.6O.00.01 to CB].
Acknowledgements
We acknowledge Parques Nacionales (Ministerio para la Transicion Ecologica y Reto Demografico, Spain) and Generalitat Valenciana (Regional Government of Valencia, Spain) for the support of the Montgo-Denia Research Station. Editing was provided by Sea Pen Scientific Writing.

### Author Declarations

University of Alicante

### Summary of Updates

The most relevant changes in the manuscript are: (1) As suggested by Reviewer 1, (Lewin 1986; Perez, Monsalve, and Marquez 2015) we have updated our analysis. In the original version of the manuscript, time series of inferred infection ended on 28 April 2020, when the first phase of the National Seroprevalence Study was concluded. Now, we have updated them to 29 November 2020, when the last phase of the same study was finished. That is an extension from 4 to 11 months. REMEDID depends on seroprevalence studies since they are needed to estimate the Case Fatality Ratio (CFR). The extension of time series had a great impact on the results of the study, since new information related to the second wave arose and had to be analyzed. That is the reason for so much changes in the data and result sections. However, the methodology and the discussion are quite similar to the original version. (2) As suggested by the Editorial Board Member, we have included a validation test for REMEDID as a new Appendix. Validation of REMEDID results presented in the manuscript is quite complicated due to the lack of accurate data to compare with. Neither dynamical models could be used since they also need accurate data to be tuned. Thus, we propose a proof of concept based in a modification of REMEDID to make it comparable with available official data. The results are positive.

