## Supplementary material - REMEDID MATLAB code for "Retrospective Methodology to Estimate Daily Infections from Deaths (REMEDID) in COVID-19: the Spain case study": Supplementary Garcia et al REMEDID MATLAB.m

```
%-------------------------------------------------------------------------
%-------------------------------------------------------------------------
%-------------------------------------------------------------------------
%
% This function implements de REMEDID algorithm
% REMEDID: Retrospective Methodology to Estimate Daily Infections from 
%          Deaths  
%
% Written by 
%                       David Garcia-Garcia
%                       University of Alicante, Spain
%                      
%
%                                                                June 2020
%-------------------------------------------------------------------------
%-------------------------------------------------------------------------
%-------------------------------------------------------------------------
% 
% This algorithm has been designed to estimate infections during the 
% COVID-19 pandemic, although it can be used for any disease causing death.
%
% This function is freely distributed without any warranty. It has been
% tested for Matlab R2019b
%
%-------------------------------------------------------------------------
% This function should be cited as:
% Garcia-Garcia, D., I. Vigo, E. S. Fonfria, Z. Herrador, M. Navarro, and
% C. Bordehore. Retrospective Methodology to Estimate Daily Infections from
% Deaths (REMEDID) in COVID-19: the Spain case study. XXXXX, XXX (2020).
%-------------------------------------------------------------------------
%
% INPUTS:
%   - deaths: time series of daily deaths. It must be a row or a column.
%   - Incubation period (IP) distribution:
%       - IP_distribution: 'Gamma', 'Lognormal', 'Normal', 'Weibull'
%         (Matlab supports more distibution, but these are the most usual 
%          for this study)
%       - IP_parameter_1
%       - IP_parameter_2
%   - Illness onset to death (IOD)  distribution:
%       - IOD_distribution: 'Gamma', 'Lognormal', 'Normal', 'Weibull'
%         (Matlab supports more distibution, but these are the most usual 
%          for this study)
%       - IOD_parameter_1
%       - IOD_parameter_2
%   - min_percentage: Infections close to the end of the time series are 
%                     inferred only with a percentage of their associated 
%                     deaths. The infection time series is truncated in 
%                     such way that infections are estimated at least with 
%                     the minimum of percentage defined by the parameter. 
%                     Usually, min_percentage = 95 (%)
%   - CFR: Case Fatality Ratio in percentage.
%   - plot_option: If plot_option=1, the probability distribution functions 
%                  are plotted
%
% Parameters 1 and 2 are different for each distribution. See Matlab 
% documentation for further details. In Matlab R2019b:
% - Gamma: 
%       - Parameter_1: Shape parameter
%       - Parameter_2: Scale parameter
% - Lognormal: 
%       - Parameter_1: Mean of logarithmic values
%       - Parameter_2: Standard deviation of logarithmic values
% - Normal: 
%       - Parameter_1: Mean 
%       - Parameter_2: Standard deviation 
% - Weibull: 
%       - Parameter_1: Scale parameter
%       - Parameter_2: Shape parameter
%       
%
% OUTPUTS:
%   - infections: time series of daily infections
%   - Nmin: Last Nmin days of the "infections" time series must be 
%           eliminated to accomplish with min_percentage  
%   - Nextra: "infections" time series is longer than "deaths" time series
%             in "Nextra" days.
%             Nextra is the number of days from first infection to first 
%             day in "deaths" time series. 


function [infections, Nmin, Nextra] = remedid(deaths, ...
                              IP_distribution,  IP_parameter_1,  IP_parameter_2,...
                              IOD_distribution, IOD_parameter_1, IOD_parameter_2, ...
                              min_percentage, CFR,...
                              plot_option)


%% ---------------------
% Incubation Period (IP)
%-----------------------

% Probability Density Function (PDF)
if strcmp('Gamma', IP_distribution)
    pdf_IP = makedist('Gamma','a', IP_parameter_1, 'b', IP_parameter_2  );
elseif strcmp('Lognormal', IP_distribution)
    pdf_IP = makedist('Lognormal','mu', IP_parameter_1,'sigma', IP_parameter_2  );
elseif strcmp('Normal', IP_distribution)
    pdf_IP = makedist('Normal','mu', IP_parameter_1,'sigma', IP_parameter_2  );
elseif strcmp('Weibull', IP_distribution)
    pdf_IP = makedist('Weibull','a', IP_parameter_1,'b', IP_parameter_2  );
else
    error('Check the distribution of the Incubation Period')
end

% Numerical values of the PDF
N=60;           %Days to evaluate the PDF. 60 is enough for COVID-19. You may like change it
h_step = 0.1;   %Length of step 
x_IP = 0:h_step:N;
y_IP = pdf(pdf_IP,x_IP);


%% --------------------------
% Illnes Onset to Death (IOD)
%----------------------------

% Probability Density Function (PDF)
if strcmp('Gamma', IOD_distribution)
    pdf_IOD = makedist('Gamma','a', IOD_parameter_1, 'b', IOD_parameter_2  );
elseif strcmp('Lognormal', IOD_distribution)
    pdf_IOD = makedist('Lognormal','mu', IOD_parameter_1,'sigma', IOD_parameter_2  );
elseif strcmp('Normal', IOD_distribution)
    pdf_IOD = makedist('Normal','mu', IOD_parameter_1,'sigma', IOD_parameter_2  );
elseif strcmp('Weibull', IOD_distribution)
    pdf_IOD = makedist('Weibull','a', IOD_parameter_1,'b', IOD_parameter_2  );
else
    error('Check the distribution of the Illnes Onset to Death')
end

% Numerical values of the PDF
x_IOD = 0:h_step:N;
y_IOD = pdf(pdf_IOD,x_IOD);


%% ---------------------------------------------------------
% Convolution of pdf_IP and pdf_IOD: Infection to Death (ID)
%-----------------------------------------------------------

pdf_aux = zeros(length(x_IP), 2*length(x_IP)-1 )*NaN;

for x=1:length(x_IOD)
    
    pdf_aux(x, x:x+length(x_IP)-1 ) = y_IP(x) * y_IOD * h_step;
    
end

pdf_convolution = nansum(pdf_aux, 1);
x_convolution = 0:h_step:2*N;


% Daily version: Infection to Death (ID)
x = 0:N;
k=0;
for i=1:1/h_step:length(pdf_convolution)-1
    k=k+1;
    pdf_convolution_daily(k) = mean(pdf_convolution(i:i+9));
end

pdf_ID = pdf_convolution_daily(1: length(x));


%% --------------
% Plot (optional)
% ---------------

% It is plot only if plot_option==1
if plot_option==1
    
    figure('Renderer', 'painters', 'Position', [10 10 700 350])
    
    hold on, grid on
    h_IP  = plot(x_IP,  y_IP,  'linewidth',2);
    h_IOD = plot(x_IOD, y_IOD, 'linewidth',2);
    h_convolution = plot(x_convolution, pdf_convolution,'linewidth',2);
    
    
    legend([h_IP, h_IOD, h_convolution],...
        ['Incubation period '],...
        ['Illnes onset to death '],...
        ['Infection to death '],...
        'location', 'northeast') ;
    
    xlim([0,N])
    xlabel('Days ','FontSize',15)
    
    aa = get(gca,'XTickLabel');
    set(gca,'XTickLabel',aa,'fontsize',13)
    
    title('Probability Density Functions ', 'fontweight','bold','FontSize',25);
    
end


%% ---------------------------------------------------------
% Cumulative Distribution Function (CDF) of pdf_convolution
%-----------------------------------------------------------

for i=1:length(pdf_ID)
    cdf_ID(i) = sum(pdf_ID(1:i));
end

ind = find(cdf_ID < min_percentage/100 );

%Minimal number of days needed accordingly to min_percentage
Nmin = ind(end);


%% ----------
% Infections 
%------------

% This number must be larger than the number of days between the first day 
% of inferred infections and the first day of deaths time series. It will
% be adjust later.
Nextra_aux = 200;

%Check dimension of deaths:
[rows,colu] = size(deaths);
if     rows==1  
elseif colu==1, deaths = deaths';
else, error('Check dimensions of deaths time series')
end

% Nextra zeros are added before the beginning of deaths time series
deaths_extended = [zeros(1, Nextra_aux), deaths];

         
Nextended = length(deaths_extended);
infections_CFR100 = zeros(1, Nextended);

for i=1:Nextended
    
    aux = deaths_extended(i:end);
    
    if length(aux)>N+1
        k = length(aux) - (N+1);
        aux_pd = [pdf_ID, zeros(1,k)];
    else
        aux_pd = pdf_ID(1:length(aux));
    end
    
    % Equation 1:
    infections_CFR100(i) = sum(  aux .* aux_pd  );
    
end
    


% Equation 2:
infections_aux = round( infections_CFR100 /CFR *100 );


% Time series is cut to avoid zeros at the begining:
ind_positive = find(infections_aux > 0);
infections = infections_aux(ind_positive);

Nextra = length(infections) - length(deaths);
```
